# Immuno-epidemiological life-history and the dynamics of SARS-CoV-2 over the next five years

**DOI:** 10.1101/2020.07.15.20154401

**Authors:** Chadi M. Saad-Roy, Caroline E. Wagner, Rachel E. Baker, Sinead E. Morris, Jeremy Farrar, Andrea L. Graham, Simon A. Levin, C. Jessica E. Metcalf, Bryan T. Grenfell

**Affiliations:** Lewis-Sigler Institute for Integrative Genomics, Princeton University, Princeton NJ 08540, USA; Department of Ecology and Evolutionary Biology, Princeton University, Princeton NJ 08544, USA; Princeton Environmental Institute, Princeton University, Princeton NJ 08544, USA; Department of Pathology and Cell Biology, Columbia University Medical Center, New York NY 10032, USA; The Wellcome Trust, London, UK; Princeton School of Public and International Affairs, Princeton University, Princeton NJ 08544, USA; Fogarty International Center, National Institutes of Health, Bethesda MD 20892, USA

## Abstract

Uncertainty in the immune response to SARS-CoV-2 may have implications for future outbreaks. We use simple epidemiological models to explore estimates for the magnitude and timing of future Covid-19 cases given different impacts of the adaptive immune response to SARS-CoV-2 as well as its interaction with vaccines and nonpharmaceutical interventions. We find that variations in the immune response to primary SARS-CoV-2 infections and a potential vaccine can lead to dramatically different immunity landscapes and burdens of critically severe cases, ranging from sustained epidemics to near elimination. Our findings illustrate likely complexities in future Covid-19 dynamics, and highlight the importance of immunological characterization be-yond the measurement of active infections for adequately characterizing the immune landscape generated by SARS-CoV-2 infections.

## Introduction

The current Covid-19 pandemic caused by the novel SARS-CoV-2 betacoronavirus *(β*–CoV) has resulted in substantial morbidity and mortality, with over ten million confirmed cases world-wide at the time of writing. In order to curb viral transmission, non-pharmaceutical interventions (NPIs) including business and school closures, restrictions on movement, and total lock-downs have been implemented to various degrees around the world. Major efforts to develop effective vaccines and antivirals are ongoing.

Understanding the future trajectory of this disease requires knowledge of the population-level landscape of immunity generated by the life histories of SARS-CoV-2 infection or vaccination among individual hosts, particularly in the context of the acquisition, re-transmission, and clinical severity of secondary infections. The nature of acquired immune responses following natural infection varies substantially among pathogens. At one end of this immune spectrum, natural infection with measles *(1)* or smallpox *(2)* virus results in lifelong protection from the re-acquisition and re-transmission of secondary infections. Many other infections (e.g. influenza *(3)*, RSV *(4)*) confer imperfect or transient clinical and transmission-blocking immunity. Finally, phenomena such as antibody-dependent enhancement (ADE) associated with prior natural infection (e.g. dengue *(5)*) or a vaccine (e.g. RSV *(6)*) could result in more clinically severe secondary infections. Furthermore, the immunity conferred by vaccines to various pathogens rarely provides complete protection against reinfection and/or disease *(9)*, and this protection may be inferior to that acquired following natural infection *(10)*. Nevertheless, imperfect vaccines that reduce both the clinical severity and transmissibility of subsequent infections (if they do occur) can still provide population-level disease protection *(9, 11)*.

In the case of SARS-CoV-2, the nature of the immune response following natural infection remains uncertain, although efforts to measure the kinetic adaptive immune response are ongoing *(12–16)*. One approach for bounding estimates for this immune response is to examine related viruses that also belong to the *β*–CoV genus. This genus comprises other viruses that cause severe infections in humans including the Middle East Respiratory Syndrome (MERS) and SARS-CoV-1 coronaviruses *(7)* as well as the seasonal coronaviruses HCoV-HKU1 and HCoV-OC43, which are the second leading causes of the common cold *(7)*. While immunity to SARS-CoV-1 is believed to last up to 2-3 years *(17, 18)*, serum antibody levels against HCoV-OC43 have been found to wane on the time-scale of a few months *(19)* to one year *(20)* likely due in part to a combination of antigenic evolution and low antigenic dose. HCoV-HKU1 and HCoV-OC43 are thought to cause repeated infections throughout life *(21)*, though a significant biennial component in their dynamics implies at least some herd protection *(7, 8)*. Furthermore, although it is currently unclear whether ADE influences the pathogenesis of SARS-CoV-2, it has been hypothesized that severe Covid-19 cases may arise from the presence of non-neutralizing antibodies from prior coronavirus infections *(22)* following earlier proposals for related coronaviruses *(23–25)*. For example, one study found that a subset of reinfections with three endemic human coronaviruses (HCoV-NL63, HCoV-229E, and HCoV-OC43) showed enhanced viral replication during the secondary infection *(26)*. In addition to antibody-mediated immunity, T cell immunity increasingly appears to be an important component of the adaptive immune response to SARS-CoV-2; also there is evidence of T cell cross-reactivity between SARS-CoV-2 and these seasonal, endemic coronaviruses *(15, 16)*.

Various epidemiological models have been developed to capture this type of variation in immune responses on population-level infection dynamics. For instance, the well-known Susceptible-Infectious-Recovered (SIR) model is suitable for modeling the dynamics of perfectly-immunizing infections such as measles *(27)*, while the Susceptible-Infectious-Recovered-Susceptible (SIRS) model captures the epidemiology of imperfectly immunizing infections such as influenza, in which individuals eventually return to a fully or substantially susceptible class following a finite period of immunity either due to waning memory or pathogen evolution *(28)*. More complex compartmental models have also been developed to study infections characterized by intermediate immune responses lying between these two extremes such as rotavirus *(29)* and respiratory syncytial virus *(4)*.

Here we adopt a generalization of these more nuanced models, the SIR(S) model *(28)* outlined schematically in Figure 1 and Figure S1, to explore how the pandemic trajectory might unfold for different assumptions regarding the nature of the adaptive immune response to SARS-CoV-2 infection. Since different adaptive immune responses may be associated with variations in the proportion of severe secondary cases, we also consider different scenarios for this fraction in order to estimate the future clinical burden of SARS-CoV-2 infections. The model assumes different infection and immune phenotypes depending on exposure history (see Methods for the full mathematical details). Specifically, it interpolates between the fully immunizing SIR model when immunity is lifelong and the imperfectly immunizing SIRS model through the degree of susceptibility to and transmissibility of secondary infections (quantified by the parameters *ϵ* and *α*, respectively). As shown in the representative time series of Figure 1, the SIR model results in recurrent epidemics fueled by births following the pandemic peak, while the SIRS model results in shorter inter-epidemic periods due to the possibility of reinfection and the buffering of the fully susceptible birth cohort by partially immune individuals *(28)*. We begin by characterizing the effect of temporal changes in the transmission rate brought about by climate and the adoption of NPIs on the predictions of the SIR(S) model under a range of immunity assumptions. Next, we examine the effect of a transmission-reducing vaccine of varying efficacy relative to natural immunity, thus altering the relative compositions of the various immune classes. Finally, we estimate the post-pandemic immunity landscape and clinical case burden for different possible ‘futures’ shaped by the various aspects of SARS-CoV-2 biology as well as the presence or absence of these external drivers and interventions.

**Figure 1:**
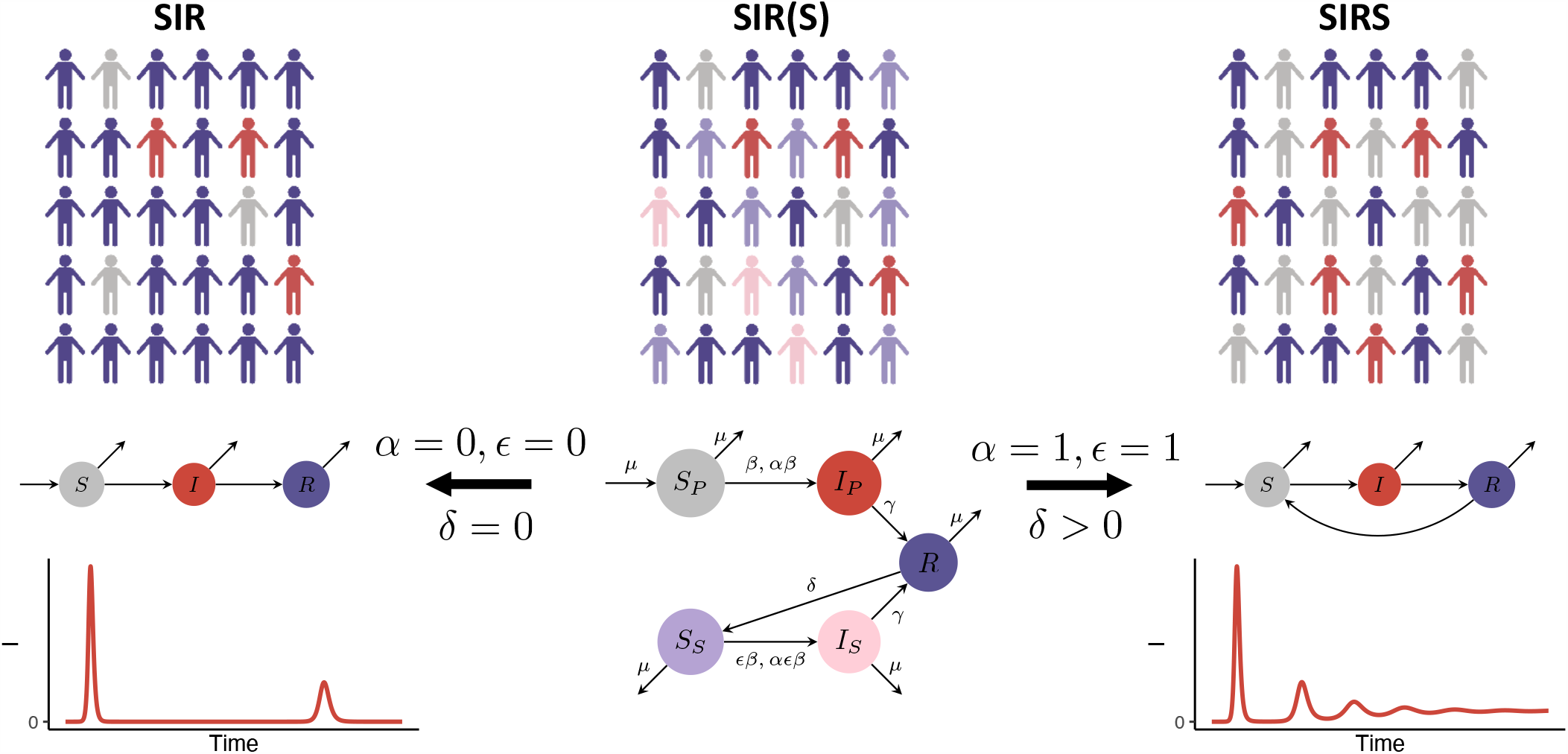
Schematic of the SIR(S) model with a flowchart depicting flows between immune classes. Here, *S*_*P*_ denotes fully susceptible individuals; *I*_*P*_ denotes individuals with primary infection that transmit at rate *β*; *R* denotes fully immune individuals (due to recovery from either primary or secondary infection), *S*_*S*_ denotes individuals whose immunity has waned at rate *δ* and are now susceptible again to infection, with relative susceptibility *ϵ*; *I*_*S*_ denotes individuals with secondary infection that transmit at a reduced rate *αβ*, and *μ* denotes the birth rate (see Methods). Illustrations and flowcharts of the limiting SIR and SIRS models are also shown (where individuals are either fully susceptible *(S)*, infected *(I)*, or fully immune *(R)*), along with a representative time series for the number of infections in each scenario. The population schematics were made through Biorender.com.

## Results and Discussion

### Seasonal transmission rates and the adoption of NPIs

In practice, during the early stages of a pandemic, reductions in transmission are nearly entirely achieved through the adoption of NPIs such as mask-wearing or social distancing. To explore the effect of the adoption of NPIs, we considered two different scenarios for timed reductions in the force of infection to 60% of its original value (in agreement with intermediate levels of social distancing in *(7)*). In Figures 2a - 2c, we plot the time series of primary and secondary infections assuming NYC climate-driven seasonality in transmission as well as single periods of NPI adoption lasting from weeks 16 to 67 (Figure 2a) or 16 to 55 (Figure 2b), and two shorter periods during weeks 16 to 55 and weeks 82 to 93 separated by normal interactions (Figure 2c). The weekly reproduction numbers corresponding to these three scenarios are plotted in Figures S2d-S2f. Although these reproduction numbers are based on those obtained for the related *β*-CoV HCoV-HKU1 and are in general lower than those estimated during the early stages of the Covid-19 pandemic *(30)*, they may be more appropriate for considering the longer-term transmission dynamics, and our results are qualitatively robust to increasing these values for *R*_0_ (results not shown).

**Figure 2:**
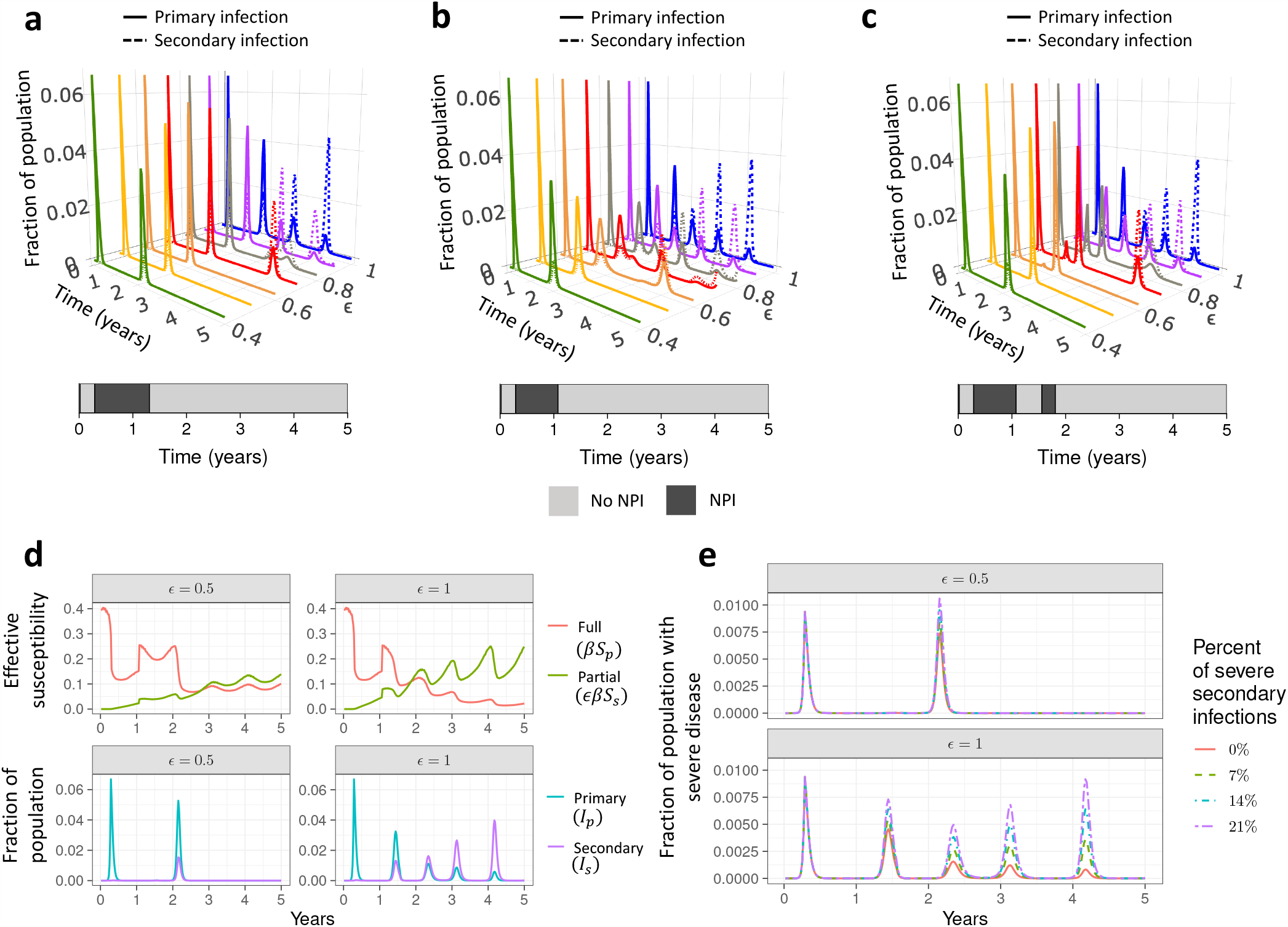
(a) to (c): Effect of NPI adoption on the time series of primary (solid lines) and secondary (dashed lines) infections with a seasonal transmission rate derived from the climate of NYC with no lag between seasonality and epidemic onset. NPIs that reduces the transmission rate to 60% of the estimated climate value are assumed to be adopted during weeks 16-67 in (a), weeks 16-55 in (b), and weeks 16-55 as well as 82-93 in (c). Colours denote individual time courses for different values of *ϵ*. (d) Time series of the force of infection arising from susceptibility (top row) and the fraction of the population that is infected (bottom row) for both primary and secondary infections, for *ϵ* = 0.5 (left column) and *ϵ* = 1 (right column) for the NPI scenario outlined in (c). (e) Time series of estimated number of severe infections for the NPI scenario defined in (c) for four different estimates of the fraction of severe cases during primary infections *(x*_sev,*p*_) and secondary infections *(x*_sev,*p*_) with *ϵ* = 0.5 (top row) and *ϵ* = 1 (bottom row). These are *x*_sev,*p*_ = 0.14, *x*_sev,*s*_ =0 (solid red line), *x*_sev,*p*_ = 0.14, *x*_sev,*s*_ = 0.07 (dashed green line), *x*_sev,*p*_ = 0.14, *x*_sev,*s*_ = 0.14 (dashed-dotted blue line), and *x*_sev,*p*_ = 0.14, *x*_sev,*s*_ = 0.21 (shortdashed-longdashed purple line).

We find that decreases in the susceptibility to secondary infection *ϵ* can delay secondary peaks (compare individual time courses for different values of *ϵ* in Figures 2a–2c). However, delayed peaks may then be larger, due to susceptible accumulation and dynamical resonance. These non-monotonicities in the timing and size of secondary peaks also occur with climate-driven seasonal transmission in the absence of NPIs (see Supplementary Materials, Section S3.3). Importantly, the delay that social distancing may cause in the timing of the secondary peak can also allow for the accumulation of fully susceptible individuals. This is illustrated in the top panels of Figure 2d, where the full *(βS*_*P*_, red curve) and partial *(ϵ βS*_*S*_, green curve) effective population-level susceptibility corresponding to a reduction in susceptibility to secondary infection by 50% *(ϵ* = 0.5) (left) and with no reduction in susceptibility to secondary infection *(ϵ* = 1) (right) for the social distancing scenario outlined in Figure 2c are shown. The corresponding fraction of primary (blue) and secondary (purple) cases are presented in the bottom panels. As can be seen, when the secondary peak does occur, the decrease in susceptibility to secondary infection *(ϵ <* 1) considered in the left panels results in a greater number of primary infections during the second peak relative to the panels on the right where *ϵ* = 1.

Finally, an essential part of the planning and management of future SARS-CoV-2 infections is the ability to characterize the magnitude and timing of severe cases requiring hospitalization. In Figure 2e we consider four different plausible scenarios for the fraction of severe secondary cases *x*_sev,*s*_ (see Methods) based on the scenario depicted in Figure 2c and assuming 14% of primary cases are severe *(31)*: no severe cases associated with secondary infection *(x*_sev,*s*_ = 0, solid red line), a reduced number of severe cases with secondary infection relative to primary infection *(x*_sev,*s*_ = 0.07, dashed green line), comparable proportions of severe cases *(x*_sev,*s*_ = 0.14, dashed-dotted blue line), and a greater proportion of severe cases with secondary infection *(x*_sev,*s*_ = 0.21, shortdashed-longdashed purple line), possibly owing to phenomena such as ADE. When the assumed fraction of severe secondary infections is high, the fraction of the population with severe infections during secondary peaks exceeds that observed during the primary peak (Figure 2e). As the proportion of secondary infections increases during the later stages of the pandemic, these findings stress that clinical epidemiological studies of repeat infections will be critical for proper planning of healthcare systems.

### Vaccination

The availability of an effective vaccine would be a key intervention against SARS-CoV-2, and numerous candidates are in development *(32, 33)*. Intuitively, if the effective vaccination rate is sufficiently high, then vaccination herd immunity generated by a transmission-blocking vaccine could control or eliminate the infection. However, this becomes harder to achieve when vaccinal and natural immunity is imperfect and secondary infections occur, or when logistical or other constraints limit vaccine deployment. Although it remains uncertain whether and when a vaccine will be available, we make the relatively optimistic assumption that a transmission-reducing vaccine begins to be introduced at *t*_vax_ = 1.5 years. We also consider seasonal transmission rates as in Figure S3 and the adoption of NPIs according to the scenario described in Figure 2b. We assume that a constant proportion *υ* ranging from 0% *≤ υ ≤* 1% of the fully and partially susceptible populations *(S*_*P*_ and *S*_*S*_) is effectively vaccinated every week and acquire transmission-blocking immunity for, on average, a period 1*/δ*_vax_. For comparison, it was estimated that in response to the 2009 H1N1 pandemic, one or more doses of the monovalent vaccine were administered to 80.8 million vaccinees during October 2009 to May 2010 in the United States *(34)*, which implies a rate of vaccination coverage of about 27% after a period of 8 months for persons aged at least 6 months in the USA, although rates between different nations varied *(35)*. This crudely corresponds to a weekly vaccination rate of 1% *(36)*. Finally, we assume that the immunity conferred from effective vaccination wanes at rate *δ*_vax_ which in general may differ from the waning rate of immunity from natural infection, *δ*. The modified set of ordinary differential equations in this scenario corresponding to the flowchart in Figure S11a is presented in the Methods.

In Figure 3a we consider the long-term equilibrium infection burden (see Methods) following vaccination at a weekly rate *υ* for a variety of immunity assumptions. As expected, a reduction in the susceptibility to secondary infections *(ϵ)* results in a smaller number of infections at steady state in the absence of vaccination. Further, both *ϵ* and the duration of vaccine immunity (1*/δ*_vax_) affect the vaccination rate required to achieve a disease-free state at equilibrium. In the limit of fully immunizing primary infections and vaccines *(ϵ* = 0), relatively low vaccination rates are required to achieve zero infections at steady state. However, as immunity becomes more imperfect (larger *ϵ)*, increasingly high vaccination rates are required to eliminate infections particularly when the duration of vaccine immunity is short. This is further emphasized in Figure 3b, where the minimum vaccination rate *υ* required to achieve a disease-free state at equilibrium (see Methods) is plotted as a function of *ϵ* for different values of the duration of vaccine immunity. Altogether, this illustrates that reductions in infection achievable through vaccination are inherently related to the efficacy of the vaccine and the nature of the adaptive immune response *(37)*.

**Figure 3:**
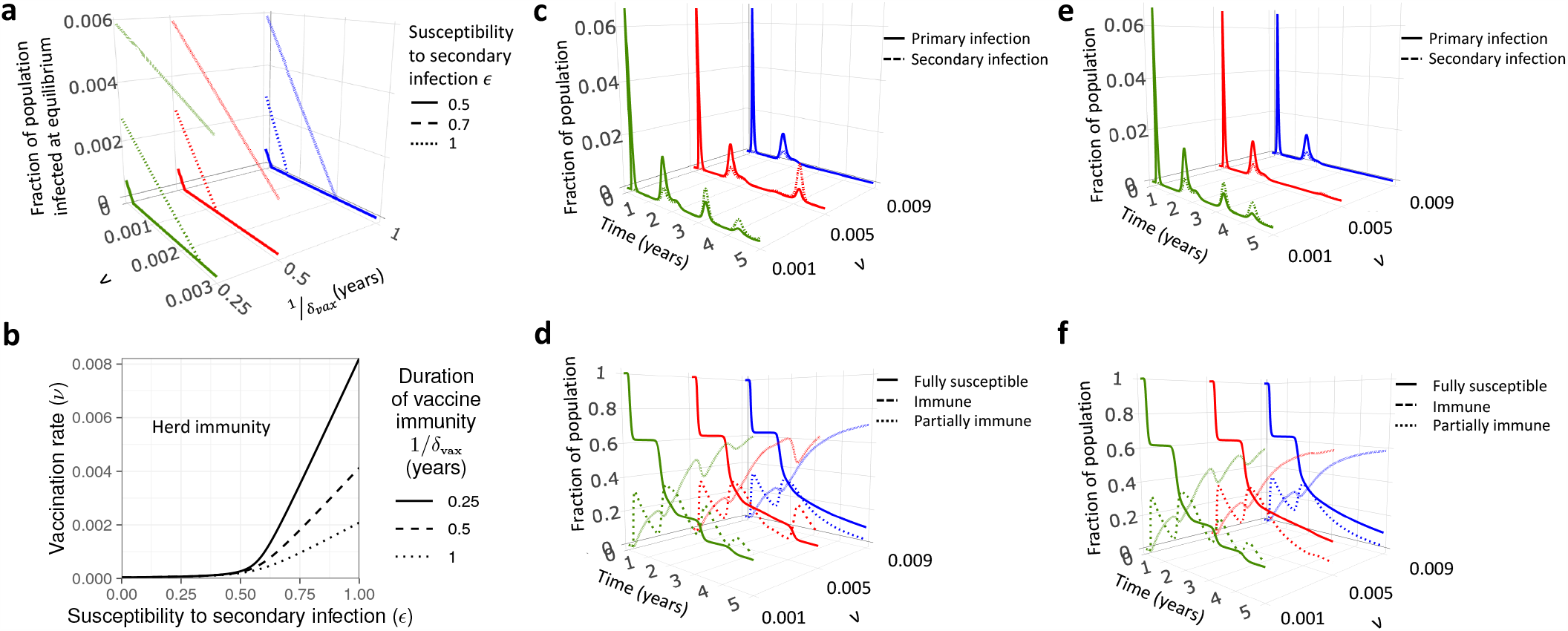
(a) Total infected fraction of the population at equilibrium as a function of the vaccination rate *υ* for different values of the duration of vaccine immunity (1*/δ*_vax_ = 0.25 years: green line, 1*/δ*_vax_ = 0.5 years: red line, and 1*/δ*_vax_ =1 year: blue line) and the susceptibility to secondary infection *(ϵ* = 0.5: solid line, *ϵ* = 0.7: dashed line, and *ϵ* = 1: dotted line). (b) Vaccination rate *υ* required to achieve a disease-free state at equilibrium as a function of *ϵ* for different values of the duration of vaccine immunity (1*/δ*_vax_ = 0.25 years: solid line, 1*/δ*_vax_ = 0.5 years: dashed line, and 1*/δ*_vax_ = 1 year: dotted line). In (a) and (b) the relative transmissibility of secondary infections and duration of natural immunity are taken to be *α* =1 and 1*/δ* = 1 year, respectively, and the transmission rate is derived from the mean value of seasonal NYC-based weekly reproduction numbers (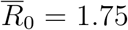, see Methods and Figure S2c). (c)-(f): Time series of the various immune classes plotted for different values of the vaccination rate *υ*. The top row ((c) and (e)) contains the time series of primary (solid lines) and secondary (dashed lines) infections, while the bottom row ((d) and (f)) contains the time series of the fully susceptible (solid lines), immune (dashed lines), and partially immune (dotted lines) subpopulations. The duration of vaccine immunity is taken to be 1*/δ*_vax_ = 0.5 years (shorter than natural immunity) in (c) and (d), and 1*/δ*_vax_ = 1 year (equal to natural immunity) in (e) and (f). The susceptibility to secondary infection, relative transmissibility of secondary infections, and duration of natural immunity are taken to be *ϵ* = 0.7, *α* = 1, and 1*/δ* =1 year, respectively. Vaccination is introduced 1.5 years after the onset of the epidemic following a 40 week period of social distancing during which the force of infection was reduced to 60% of its original value during weeks 16 to 55 (i.e. the scenario described in Figure 2b of the main text), and a seasonal transmission rate derived from the climate of NYC with no lag is assumed.

We next explore the short-term dynamical effect of vaccination in Figures 3c-3f. In Figure S11b, the ratio of the total number of primary infections during years 1.5-5 inclusive (i.e. after the vaccine is introduced) relative to the zero vaccination case for different values of the vaccination rate *υ* and the duration of vaccine immunity 1*/δ*_vax_ is shown. Figure S11c shows the equivalent for secondary infections. The burden of primary infections decreases with increasing vaccination rate for a given value of vaccine immunity 1*/δ*_vax_. However, for the shortest durations of vaccine immunity 1*/δ*_vax_, the degree of achievable reductions in the number of secondary cases begins to saturate even for large vaccination rates. This saturation is due to the rapid return of vaccinated individuals to the partially susceptible class if vaccine immunity is short-lived. Further, when the duration of vaccine immunity is very short, the total number of secondary cases can be higher when vaccination takes place for the particular model and parameters considered. To further emphasize the dependence of the model results on the vaccination rate and duration of vaccine immunity, we present time courses of infections and immunity for different durations of vaccine immunity and vaccination rates in Figures 3c - 3f as well as Figures S11d and S11e. In line with intuition, the model illustrates that both high vaccination rates and relatively long durations of vaccine-induced immunity are required to achieve the largest reductions in secondary infection burdens.

### Clinical case burden and immunity landscape for different possible futures

Figure 4 is a synoptic view of the impact of vaccination and natural immunity in four different ‘futures’ on the immune landscape and incidence of severe disease. In all four scenarios, we assume seasonal transmission (c.f. Figure S3) and social distancing according to the scenario depicted in Figure 2b. Figures 4a and 4b correspond to futures without vaccination, with Figure 4a illustrating a more pessimistic scenario of greater susceptibility to secondary infections *(ϵ* = 0.7), a relatively short period of natural immunity (1*/δ* = 0.5 years), and a greater proportion of severe cases with secondary infection possibly owing to phenomena such as ADE. In contrast, the more optimistic future of Figure 4b assumes reduced susceptibility to secondary infections *(ϵ* = 0.5), a longer duration of natural immunity (1*/δ* = 2 years), and a smaller proportion of severe cases with secondary infection. In both cases, the initial pandemic wave is the same, but in the more optimistic scenario (Figure 4b), natural immunity is longer lasting and consequently subsequent infection peaks are delayed. Furthermore, the reduction in susceptibility to secondary infection (smaller *ϵ)* in Figure 4b suppresses the later peaks dominated by secondary infections (Figure 4a) and substantially less depletion of fully susceptible individuals occurs. In Figures 4c and 4d, these pessimistic and optimistic scenarios are translated into futures with vaccination, which is assumed to be introduced at a weekly rate of *υ* = 1% at *t*_vax_ = 1.5 years. The future described in Figure 4c assumes all the same outcomes as in Figure 4a and incorporates vaccination with short-lived vaccine immunity (1*/δ*_vax_ = 0.25 years). The future presented in Figure 4d assumes all the same outcomes as in Figure 4b in addition to vaccine immunity lasting 1*/δ*_vax_ =1 year.

**Figure 4:**
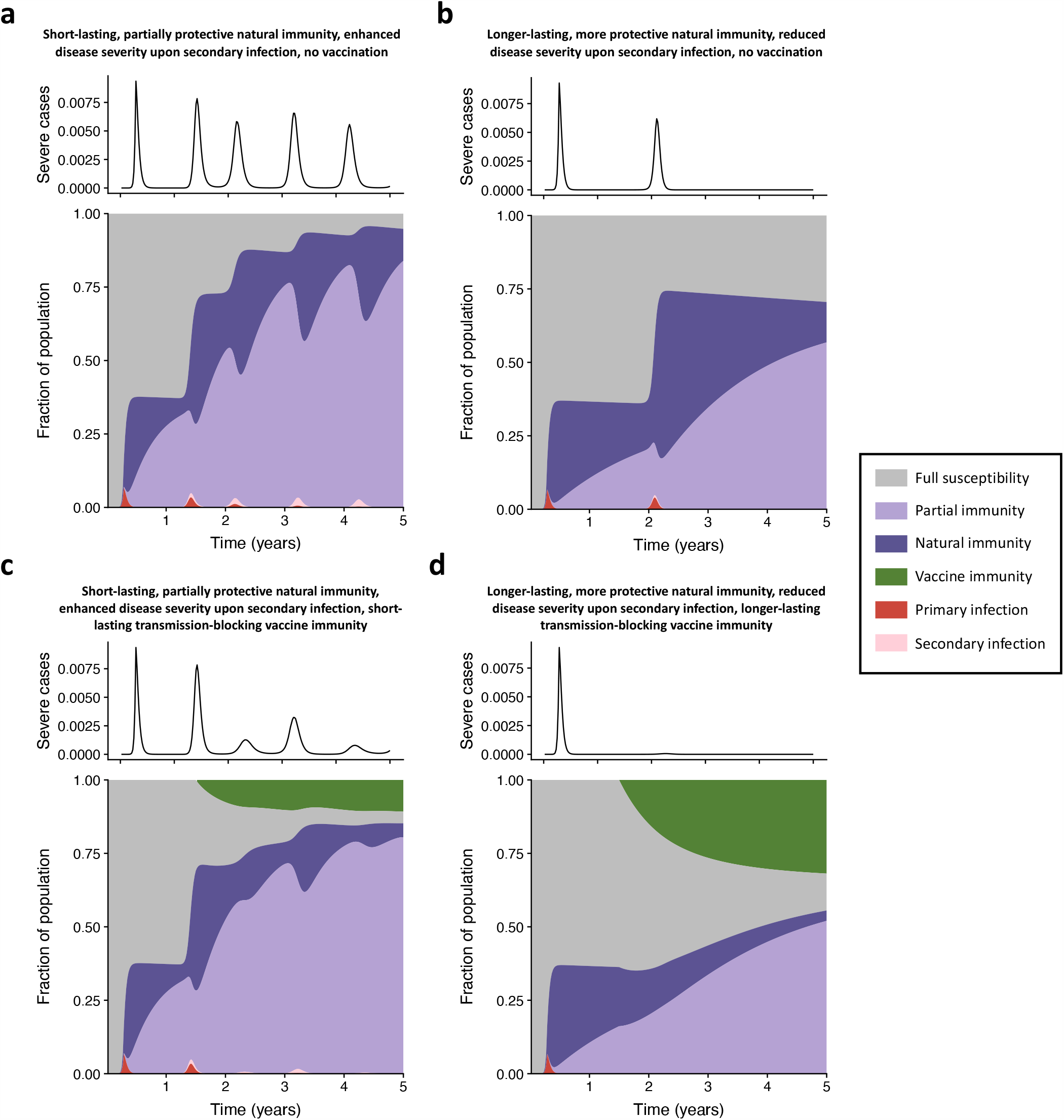
Time series of fraction of the population with severe cases *I*_sev_ (top) and area plots of the fraction of the population comprising each immune class (bottom) over a five year time period under four different future scenarios. In all plots, the relative transmissibility of secondary infections is taken to be *α* = 1, a seasonal transmission rate derived from the climate of NYC with no lag is assumed, and a period of social distancing during which the force of infection is reduced to 60% of its original value during weeks 16 to 55 (i.e. the scenario described in Figure 2b) is enforced. In (a) and (b), no vaccination occurs. (a) Corresponds to a more pessimistic natural immunity scenario with *ϵ* = 0.7, 1*/δ* = 0.5 years, and 21% of secondary cases being severe. (b) Corresponds to a more optimistic natural immunity scenario with *ϵ* = 0.5, 1*/δ* = 2 years, and 7% of secondary cases being severe. In (c) and (d), vaccination is introduced at a weekly rate of *υ* = 1% at *t*_vax_ = 1.5 years after the onset of the epidemic. (c) Corresponds to all the parameters in (b) along with vaccine immunity lasting 1*/δ*_vax_ = 0.25 years, while (d) corresponds to all the same parameters as in (b) along with vaccine immunity lasting 1*/δ*_vax_ =1 year.

Figures 4c and 4d emphasize the important role that even an imperfect vaccine could have in SARS-CoV-2 dynamics and control *(9, 11)*. With vaccination, subsequent peaks in clinically severe cases are substantially reduced, although in the pessimistic future later infection peaks dominated by secondary infections can still occur (Figure 4c). Furthermore, if a transmission-blocking vaccine confers a relatively long period of protection, and if we make optimistic assumptions regarding the nature of the adaptive immune response (Figure 4d), a sufficient proportion of fully susceptible individuals can be immunized, thus suppressing future outbreaks. These trends are qualitatively conserved for different vaccine deployment strategies, such as a pulse of immunization at *t*_vax_ = 1.5 years in which a fixed percentage of the fully and partially susceptible populations *(S*_*P*_ and *S*_*S*_) are vaccinated (see Figure S12). However, without sustained immunization strategies, the waning of vaccine immunity results in less susceptible depletion over time, and larger future outbreaks relative to the scenarios presented in Figures 4c and 4d. Overall, Figure 4 suggests that relying on the status of infection of an individual as the main observable during an ongoing epidemic is insufficient to characterize the complex immune landscape that is generated. Given the increasingly recognized importance of both T cell- *(15, 16)* and antibody-mediated *(12–14)* adaptive immune responses in the recovery from SARS-CoV-2 infection, it is very likely that more sophisticated methods including regular testing of serology and T cell immunity will be required *(38)*.

## Conclusion

We have examined how plausible variations in the natural immune response following SARS-CoV-2 infection as well as various external drivers and interventions may shape the longer-term clinical burden and immunity landscape to this disease. Here we summarize the key findings and complexities illustrated by this simple model along with caveats for their interpretation; a full summary of all caveats and future directions is given in Section S2.

In locations where we expect substantial climatically-driven seasonal variation in transmission, such as New York City, the model predicts that a reduction in susceptibility to secondary infection or a longer duration of immunity may lead to a larger secondary infection peak, and this peak may occur earlier when the duration of natural immunity is longer. In geographical areas with smaller annual fluctuations in climate, we find that this non-monotonic behaviour is increasingly suppressed, although these results are sensitive to the assumed form of climatic influences on SARS-CoV-2 transmission, which we have taken here to be very similar to those of the related *β*–CoV HCoV-HKU1. Indeed, even with the incorporation of NPIs (which in general may be adopted differently than the scenarios considered here), the clinical burden of subsequent infection peaks is sensitive to the relative fraction of primary and secondary cases as well as the fraction of severe cases for each category, which intimately depend on the nature of the adaptive immune response. In turn, the severity of an infection could affect the nature of the subsequent adaptive immune response through antigen priming or T cell exhaustion. Finally, the disease dynamics predicted by the model can be substantially altered using a highly simplified scenario for the introduction of a vaccine, although the achievable reductions in infections are strongly dependent on the efficacy of the vaccine and the nature of the adaptive immune response. Nevertheless, even with imperfect vaccine immunity, reductions in the number of primary cases through vaccination may have important clinical implications.

Altogether, this work emphasizes the complex dependence of the immune landscape generated by SARS-CoV-2 infection on the presently largely unknown nature of the adaptive immune response to this virus and the efficacy of potential future vaccines. Depending on how these unfold, the model predictions for future clinical burdens range from sustained epidemics to near case elimination. Consequently, accurately characterizing the nature of immunity to SARS-CoV-2 will be critical for the management and control of future infections.

## Data Availability

The manuscript does not include new data.

## Acknowledgements

CMSR acknowledges support from the Natural Sciences and Engineering Research Council of Canada through a Postgraduate-Doctoral Scholarship. CEW is an Open Philanthropy Project fellow of the Life Sciences Research Foundation. REB is supported by the Cooperative Institute for Modelling the Earth System (CIMES). SAL and CMSR acknowledge support from the James S. McDonnell Foundation 21st Century Science Initiative Collaborative Award in Understanding Dynamic and Multi-scale Systems. SAL acknowledges support from the C3.ai Digital Transformation Institute, the National Science Foundation under grant CNS-2027908, and the National Science Foundation Expeditions Grant CCF1917819. BTG acknowledges support from the US CDC.

